# Effectiveness of COVID-19 vaccines against the B.1.617.2 variant

**DOI:** 10.1101/2021.05.22.21257658

**Authors:** Jamie Lopez Bernal, Nick Andrews, Charlotte Gower, Eileen Gallagher, Ruth Simmons, Simon Thelwall, Julia Stowe, Elise Tessier, Natalie Groves, Gavin Dabrera, Richard Myers, Colin Campbell, Gayatri Amirthalingam, Matt Edmunds, Maria Zambon, Kevin Brown, Susan Hopkins, Meera Chand, Mary Ramsay

## Abstract

**Background:** The B.1.617.2 COVID-19 variant has contributed to the surge in cases in India and has now been detected across the globe, including a notable increase in cases in the UK. We estimate the effectiveness of the BNT162b2 and ChAdOx1 COVID-19 vaccines against this variant.

**Methods:** A test negative case control design was used to estimate the effectiveness of vaccination against symptomatic disease with both variants over the period that B.1.617.2 began circulating with cases identified based on sequencing and S-gene target status. Data on all symptomatic sequenced cases of COVID-19 in England was used to estimate the proportion of cases with B.1.617.2 compared to the predominant strain (B.1.1.7) by vaccination status.

**Results:** Effectiveness was notably lower after 1 dose of vaccine with B.1.617.2 cases 33.5% (95%CI: 20.6 to 44.3) compared to B.1.1.7 cases 51.1% (95%CI: 47.3 to 54.7) with similar results for both vaccines. With BNT162b2 2 dose effectiveness reduced from 93.4% (95%CI: 90.4 to 95.5) with B.1.1.7 to 87.9% (95%CI: 78.2 to 93.2) with B.1.617.2. With ChAdOx1 2 dose effectiveness reduced from 66.1% (95% CI: 54.0 to 75.0) with B.1.1.7 to 59.8% (95%CI: 28.9 to 77.3) with B.1.617.2. Sequenced cases detected after 1 or 2 doses of vaccination had a higher odds of infection with B.1.617.2 compared to unvaccinated cases (OR 1.40; 95%CI: 1.13-1.75).

**Conclusions:** After 2 doses of either vaccine there were only modest differences in vaccine effectiveness with the B.1.617.2 variant. Absolute differences in vaccine effectiveness were more marked with dose 1. This would support maximising vaccine uptake with two doses among vulnerable groups.

## Introduction

India has experienced a surge in COVID-19 cases since late March 2021 reaching over 400,000 cases and 4,000 deaths reported each day in early May 2021.(1) This has resulted in hospital services becoming overwhelmed and scarcity in oxygen supplies (2) While only a small proportion of samples have been sequenced, B.1.617 lineages have dominated. B.1.617.2 was first detected in India in December 2020 and became the most commonly reported variant in the country from mid-April 2021.(1) As of 19 of May 2021, the variant had been detected in 43 countries across 6 continents in GISAID.(3) In the UK, there has been a rapid increase in cases with this variant associated with travel to India and community transmission. (4)

The UK has achieved rapid rollout of COVID-19 vaccines, starting with deployment of the Pfizer-BioNTech BNT162b2 mRNA vaccine from December 2020, followed soon afterwards by the Oxford-Astrazeneca ChAdOx1 adenovirus vector vaccine from January 2021, and more recently the Moderna mRNA-1273 vaccine. The vaccine was initially prioritised for older adults, carers and health and social care workers, with subsequent rollout to those in clinical risk groups and younger age cohorts.(5) At an early stage of the rollout, and following advice from the Joint Committee of Vaccines and Immunisation, a policy decision was made to use an extended dosing interval of up to 12 weeks, in order to maximise the number of vulnerable individuals receiving the first dose during the second wave of the pandemic.(6)

Evidence from clinical trials has found the vaccines to be highly efficacious at preventing symptomatic disease.(7-9) This has been backed by real world evidence showing high levels of effectiveness against symptomatic disease, infection and severe disease.(10-14) The early findings from clinical trials were primarily undertaken in settings where the original Wuhan SARS-CoV-2 strain was the main circulating virus and data on effectiveness against different variants is limited. The B.1.1.7 variant, was first identified in the UK, and was the predominant lineage seen between January and May 2021. There is now substantial evidence that levels of protection conferred by vaccination are similar to those observed in the clinical trials, with additional protection against severe disease.(10, 11, 15-17) Laboratory data indicates that the B.1.351 variant has reduced neutralization by sera from vaccinated individuals.(18, 19) Observational data from Qatar indicated modestly reduced effectiveness against symptomatic disease caused by this variant but high levels of effectiveness against severe, critical or fatal disease in those vaccinated with BNT162b2.(17) Furthermore a trial of the Novavax NVX-CoV2373 vaccine found 51.0% efficacy against B.1.351.(20) Finally, high levels of neutralisation have been seen with the P1 variant with BNT162b2 elicited sera, though field measurements of vaccine effectiveness have not been reported.(19, 21)

The B.1.617.2 variant is characterised by spike protein mutations T19R, Δ157-158, L452R, T478K, D614G, P681R, and D950N.(1) Several of these mutations may impact on immune responses directed towards the key antigenic regions of receptor binding protein (452 and 478) and deletion of part of the N terminal domain.(22) P681R is at the S1/S2 cleavage site and studies have suggested that strains with mutations at that site may have increased replication, leading to higher viral loads and increased transmission.(23) Immunogenicity data have not yet been reported for the B.1.617.2 variant. Furthermore, no data have been reported on the effectiveness of COVID-19 vaccines against clinical outcomes with this variant.

In this study we aim to estimate the effectiveness of COVID-19 vaccines against symptomatic disease with the B.1.617.2 variant.

## Methods

### Study design

Two approaches were used to estimate the effect of vaccination on the B.1.617.2 variant:

First, a test negative case control (TNCC) design was used to estimate vaccine effectiveness against symptomatic disease with the B.1.617.2 variant compared to the B.1.1.7 variant over the same period. This approach has been described in detail elsewhere.(10) Briefly vaccination status is compared in symptomatic cases to those who report symptoms but test negative. This helps to control for biases related to health seeking behaviour, access to testing and case ascertainment.

Second, the proportion of cases with the B.1.617.2 variant relative to the main circulating virus (the B.1.1.7 variant) was estimated by vaccination status. The underlying assumption was that if the vaccine is equally effective against each variant a similar proportion of cases with either variant would be expected in unvaccinated compared to vaccinated individuals. Conversely if the vaccine is less effective against B.1.617.2, the variant would be expected to make up a higher proportion of cases more than three weeks after vaccination, when compared to unvaccinated individuals.

### Data sources

#### Vaccination status

Data on all individuals in England vaccinated with COVID-19 vaccines is available in a national vaccination register (the National Immunisation Management System, NIMS). Data, including date of each dose of vaccine and the vaccine type, were extracted on 17 May 2021 with vaccinations to 16 May 2021. Vaccination status was considered as dose 1 for symptom onset 21 days or more after the first dose up to the day before the 2^nd^ dose was received; dose 2 for symptom onset= 14 days or more after the second dose; and dose 1 or 2 as 21 days or more after dose 1 (including any period after dose 2).

#### COVID-19 testing

COVID-19 PCR testing in the UK is undertaken through hospital and public health laboratories, as well as community testing through drive through or home testing which is available to anyone with symptoms consistent with COVID-19 (high temperature, new continuous cough, loss or change in sense of smell or taste). All positive PCR tests between 26 October 2020 and 16 May 2021 were extracted. All negative community tests among individuals reporting symptoms were also extracted for the test negative case control analysis. Children aged under 16 years as of March 21^st^ 2021 were excluded. Data were restricted to individuals reporting symptoms and only individuals tested within 10 days of symptom onset were included to account for reduced sensitivity of PCR testing beyond this period.(24)

#### Identification of variant

Whole genome sequencing was used to identify the B.1.617.2 and B.1.1.7 variants. There has been a steady increase in the proportion of all positive samples that are sequenced, from approximately 10% in February 2021, to approximately 60% in May 2021.(4) Sequencing is undertaken at a network of laboratories including a high proportion at the Wellcome Sanger Institute, and sequences are assigned to Public Health England’s single nucleotide polymorphism based variant definitions.(25)

As a second approach for identifying each variant, laboratories using a three target PCR assay (TaqPath; Thermo Fisher) to differentiate samples testing positive or negative on the spike gene target. The B.1.1.7 variant accounts for between 98 and 100% of spike gene target failures in England. Among sequenced samples that tested S gene positive 72.2% were B.1.617.2 in April 2021 and 93.0% were B.1.617.2 in May (as of 12 May 2021).(4)

#### Data linkage

The three data sources described were linked using National Health Service number, date of birth, surname, first name, postcode and specimen identifiers and sample dates.

#### Covariates

A range of covariates that may be associated with the likelihood of being offered or accepting a vaccine and the risk of exposure to COVID-19 or specifically to either of the variants analysed, were also extracted from the NIMS and the testing data. This included age (in ten year age groups), sex, index of multiple deprivation (quintiles), ethnicity, care home status, history of foreign travel, region, period (calendar week), health and social care worker status, clinically extremely vulnerable and for the TNCC analysis, history of infection prior to the start of the vaccination programme. Individuals were considered to have travelled if they reported having travelled outside the UK and Ireland within the preceding 14 days at the point of requesting a test or if they were tested in a quarantine hotel or while quarantining at home. Postcodes were used to determine the index of multiple deprivation and unique property reference numbers were used to identify care homes.(26)

### Statistical analysis

For the TNCC analysis, logistic regression was used to estimate the odds of vaccination among symptomatic PCR confirmed cases compared to test negative controls. Cases were identified as B.1.617.2 through sequencing or if they were S-gene target positive on the TaqPath PCR assay. Cases were identified as B.1.1.7 on sequencing or if they were S-gene target negative on the TaqPath PCR assay.

If individuals had tested positive on multiple occasions within a 90 day period (which may represent a single illness episode), only the first positive test was included. A maximum of 3 randomly chosen negative test results were included per person. Tests taken within three weeks before a positive result, or after a positive result, which are more likely to be false negatives, or taken within seven days of a previous negative sample were excluded. Individuals who had previously tested positive prior to the analysis period were also excluded in order to estimate vaccine effectiveness in fully susceptible individuals. All covariates were included in the model as with previous TNCC analyses, with week included as a factor and without an interaction with region.

For the S-gene target analysis, only individuals who had tested positive on the other two PCR gene targets were included. These were restricted to week commencing 12^th^ April 2021 onwards to aim for high specificity of S-gene target positive for the B.1.617.2 variant.(4)

Vaccine effects for dose one were estimated for symptom onset dates 21 days or more after the first dose and for dose two for symptom onset dates 14 days or more after the second dose. Comparison was made to unvaccinated individuals and comparing to days 4-13 after vaccination to help account for differences in underlying risk of infection. The 0-3 day period was excluded as reactogenicity to the vaccine can cause an increase in testing which biases results as previously described.(10)

For the second analysis data on all positive samples that had been whole genome sequenced were used. The data were restricted to the period since at least 10 case of B.1.617.2 were detected per week (week commencing 5^th^ April 2021 onwards). The proportion of cases with B.1.617.2 relative to B.1.1.7 was calculated by vaccination status. Logistic regression was used to estimate the odds of testing positive with B.1.617.2 in vaccinated compared to unvaccinated individuals. The following covariates were included in the model irrespective of confounding: week of test and region (as a linear interaction), history of travel, ethnicity, age, sex, and clinically extremely vulnerable. Care home resident and deprivation were investigated for confounding and not included as odds ratio did not change (more than 1%). Samples were dropped from the analysis if they were repeats of the same variant within the same individual, if different variants were detected in the same individual within a 14 day period, and if the individual had received a mixed vaccination schedule (with two different vaccines) or had received two doses less than 19 days apart.

A sensitivity analysis was also undertaken comparing to the 0-13 day period post dose 1 (a period during which immune response to the vaccine would not be anticipated).(10) This was to control for possible unmeasured confounders which may be associated with both the likelihood of being vaccinated and the likelihood of being exposed to a variant. A further sensitivity analysis was conducted matching cases of B.1.617.2 to cases of B.1.1.7 on ethnicity, region, age group and week of sample, multiple matched controls per case were allowed.

## Results

A total of 12,675 sequenced cases were included in the analysis of which 11,621 had B.1.1.7 detected and 1,054 had B.1.617.2 detected. Characteristics of cases by variant are shown in table 1. Key differences with the B.1.617.2 variant include a higher proportion with a history of foreign travel, a higher proportion of cases in the most recent weeks (calendar weeks 17 and 18), a higher proportion of females and a higher proportion of cases in the North West region and in London, and a higher proportion in the ‘Indian or Indian British’ or ‘Any Other Asian background’ ethnic groups.

**Table 1:** characteristics by variant.

|  |  | B.1.1.7 |  |  | B.1.617.2 |  |  | Total |  |
| --- | --- | --- | --- | --- | --- | --- | --- | --- | --- |
|  |  | N | % |  | N | % |  | N | % |
| Total (row %) |  | 11,621 | 91.7% |  | 1,054 | 8.3% |  | 12,675 | 100% |
| Age | 16-29 | 4,010 | 34.5% |  | 372 | 35.3% |  | 4,382 | 34.6% |
|  | 30-39 | 3,281 | 28.2% |  | 264 | 25.0% |  | 3,545 | 28.0% |
|  | 40-49 | 2,360 | 20.3% |  | 216 | 20.5% |  | 2,576 | 20.3% |
|  | 50-59 | 1,240 | 10.7% |  | 130 | 12.3% |  | 1,370 | 10.8% |
|  | 60-69 | 551 | 4.7% |  | 45 | 4.3% |  | 596 | 4.7% |
|  | 70-79 | 140 | 1.2% |  | 22 | 2.1% |  | 162 | 1.3% |
|  | 80+ | 39 | 0.3% |  | 5 | 0.5% |  | 44 | 0.3% |
| History of travel | No | 11,460 | 98.6% |  | 1,015 | 96.3% |  | 12,475 | 98.4% |
|  | Yes | 91 | 0.8% |  | 35 | 3.3% |  | 126 | 1.0% |
|  | Unknown | 70 | 0.6% |  | 4 | 0.4% |  | 74 | 0.6% |
| Week of sample | 14 | 3,317 | 28.5% |  | 18 | 1.7% |  | 3,335 | 26.3% |
|  | 15 | 2,778 | 23.9% |  | 53 | 5.0% |  | 2,831 | 22.3% |
|  | 16 | 2,397 | 20.6% |  | 165 | 15.7% |  | 2,562 | 20.2% |
|  | 17 | 1,966 | 16.9% |  | 363 | 34.4% |  | 2,329 | 18.4% |
|  | 18 | 1,163 | 10.0% |  | 455 | 43.2% |  | 1,618 | 12.8% |
| Week of symptom onset | 12 | 27 | 0.2% |  | 0 | 0.0% |  | 27 | 0.2% |
|  | 13 | 1,429 | 12.3% |  | 8 | 0.8% |  | 1,437 | 11.3% |
|  | 14 | 3,091 | 26.6% |  | 25 | 2.4% |  | 3,116 | 24.6% |
|  | 15 | 2,542 | 21.9% |  | 89 | 8.4% |  | 2,631 | 20.8% |
|  | 16 | 2,228 | 19.2% |  | 212 | 20.1% |  | 2,440 | 19.3% |
|  | 17 | 1,609 | 13.8% |  | 442 | 41.9% |  | 2,051 | 16.2% |
|  | 18 | 695 | 6.0% |  | 278 | 26.4% |  | 973 | 7.7% |
| Sex | Female | 5,998 | 51.6% |  | 474 | 45.0% |  | 6,472 | 51.1% |
|  | Male | 5,620 | 48.4% |  | 580 | 55.0% |  | 6,200 | 48.9% |
| IMD | 1 | 3,769 | 32.4% |  | 311 | 29.5% |  | 4,080 | 32.2% |
|  | 2 | 2,567 | 22.1% |  | 228 | 21.6% |  | 2,795 | 22.1% |
|  | 3 | 2,023 | 17.4% |  | 190 | 18.0% |  | 2,213 | 17.5% |
|  | 4 | 1,798 | 15.5% |  | 196 | 18.6% |  | 1,994 | 15.7% |
|  | 5 | 1,446 | 12.4% |  | 126 | 12.0% |  | 1,572 | 12.4% |
| CEV | No | 11,396 | 98.1% |  | 1,037 | 98.4% |  | 12,433 | 98.1% |
|  | Yes | 225 | 1.9% |  | 17 | 1.6% |  | 242 | 1.9% |
| Carehome resident | No | 11,612 | 99.9% |  | 1,054 | 100.0% |  | 12,666 | 99.9% |
|  | Yes | 9 | 0.1% |  | 0 | 0.0% |  | 9 | 0.1% |
| Health or social care | No | 11,438 | 98.4% |  | 1,042 | 98.9% |  | 12,480 | 98.5% |
|  | Yes | 183 | 1.6% |  | 12 | 1.1% |  | 195 | 1.5% |
| Region | East Midlands | 1,395 | 12.0% |  | 128 | 12.1% |  | 1,523 | 12.0% |
|  | East of England | 886 | 7.6% |  | 116 | 11.0% |  | 1,002 | 7.9% |
|  | London | 832 | 7.2% |  | 211 | 20.0% |  | 1,043 | 8.2% |
|  | North East | 749 | 6.4% |  | 17 | 1.6% |  | 766 | 6.0% |
|  | North West | 2,032 | 17.5% |  | 443 | 42.0% |  | 2,475 | 19.5% |
|  | South East | 707 | 6.1% |  | 56 | 5.3% |  | 763 | 6.0% |
|  | South West | 148 | 1.3% |  | 13 | 1.2% |  | 161 | 1.3% |
|  | West Midlands | 1,232 | 10.6% |  | 51 | 4.8% |  | 1,283 | 10.1% |
|  | Yorkshire and Humber | 3,640 | 31.3% |  | 19 | 1.8% |  | 3,659 | 28.9% |
| Ethnicity | Any other Asian background | 220 | 1.9% |  | 60 | 5.7% |  | 280 | 2.2% |
|  | Any other ethnic group | 341 | 2.9% |  | 27 | 2.6% |  | 368 | 2.9% |
|  | Bangladeshi or Britis | 175 | 1.5% |  | 15 | 1.4% |  | 190 | 1.5% |
|  | Black African/Caribbean | 213 | 1.8% |  | 34 | 3.2% |  | 247 | 1.9% |
|  | Chinese | 45 | 0.4% |  | 4 | 0.4% |  | 49 | 0.4% |
|  | Indian or British Indian | 381 | 3.3% |  | 276 | 26.2% |  | 657 | 5.2% |
|  | Mixed | 181 | 1.6% |  | 19 | 1.8% |  | 200 | 1.6% |
|  | Pakistani or British | 822 | 7.1% |  | 62 | 5.9% |  | 884 | 7.0% |
|  | White | 7,524 | 64.7% |  | 399 | 37.9% |  | 7,923 | 62.5% |
|  | missing | 1,719 | 14.8% |  | 158 | 15.0% |  | 1,877 | 14.8% |

Week is 2021 calendar week; IMD = Index of Multiple Deprivation (1= least deprived, 5 = most deprived); CEV = in a clinically extremely vulnerable group

Among sequenced samples that were originally tested using the TaqPath assay, there was a high correlation between S-gene target status and the two variants under investigation with 87.5% of S-gene positive cases identified as B.1.617.2 and 99.7% of S-gene target negative cases identified as B.1.1.7 (supplementary tables 1 and 2). Results of the TNCC analysis are shown in table 2. In the ‘any vaccine’ analysis, effectiveness was notably lower after 1 dose of vaccine with B.1.617.2 cases 33.5% (95%CI: 20.6 to 44.3) compared to B.1.1.7 cases 51.1% (95%CI: 47.3 to 54.7). Results for dose 1 were similar for both vaccines. Following dose 2, the reduction in vaccine effectiveness was much smaller and non-significant: 86.8% (95%CI: 83.1 to 89.6) with B.1.1.7 and 80.9 (70.7 to 87.6) with B.1.617.2. With BNT162b2 there was a small reduction in effectiveness post dose 2 from 93.4% (95%CI: 90.4 to 95.5) with B.1.1.7 to 87.9% (95%CI:78.2 to 93.2) with B.1.617.2. Numbers vaccinated with 2 doses of ChAdOx1 were smaller and the overall 2 dose vaccine effectiveness was lower than with BNT162b2 however the difference in vaccine effectiveness between B.1.1.7 and B.1.617.2 was small and non-significant: 66.1% (95% CI: 54.0 to 75.0) and 59.8% (95%CI: 28.9 to 77.3) respectively.

**Table 2:** Vaccine effectiveness against S-gene target negative (B.1.1.7) and S-gene target positive (B.1.617.2)

| Vaccination status | Test negative controls | B.1.1.7 or S-gene target negative |  |  | B.1.617.2 or S-gene target positive |  |  |
| --- | --- | --- | --- | --- | --- | --- | --- |
|  |  | cases | cases:controls | aVE(%) | cases | cases:controls | aVE(%) |
| Unvaccinated | 58253 | 4891 | 0.084 | base | 695 | 0.012 | base |
| Any vaccine |  |  |  |  |  |  |  |
| Dose 1 | 32703 | 1481 | 0.045 | 51.1 (47.3 to 54.7) | 279 | 0.009 | 33.5 (20.6 to 44.3) |
| Dose 2 | 8483 | 74 | 0.009 | 86.8 (83.1 to 89.6) | 27 | 0.003 | 80.9 (70.7 to 87.6) |
| BNT162b2 |  |  |  |  |  |  |  |
| Dose 1 | 7036 | 344 | 0.049 | 49.2 (42.6 to 55.0) | 49 | 0.007 | 33.2 (8.3 to 51.4) |
| Dose 2 | 6412 | 28 | 0.004 | 93.4 (90.4 to 95.5) | 13 | 0.002 | 87.9 (78.2 to 93.2) |
| ChAdOx1 |  |  |  |  |  |  |  |
| Dose 1 | 25667 | 1137 | 0.044 | 51.4 (47.3 to 55.2) | 230 | 0.009 | 32.9 (19.3 to 44.3) |
| Dose 2 | 2071 | 46 | 0.022 | 66.1 (54.0 to 75.0) | 14 | 0.007 | 59.8 (28.9 to 77.3) |

Table 3 shows the adjusted odds ratios for detection of B.1.617.2 relative to B.1.1.7 in vaccinated compared to unvaccinated individuals odds of cases having B.1.617.2 detected in vaccinated individuals was higher than in unvaccinated individuals for dose 1 of any vaccine (OR 1.38; 95% CI 1.10-1.72) and dose 2 of any vaccine (OR 1.60; 0.87-2.97). Given that vaccine effectiveness against symptomatic disease with B.1.1.7 is estimated at approximately 60% after dose 1 and 85% after dose 2,(10, 27) these results would indicate effectiveness of 45% and 76% respectively for B.1.617.2. By vaccine type the reduction in vaccine effectiveness appeared to be greater with ChAdOx1 (OR 1.48; 95%CI 1.18-1.87) than BNT162b2 (OR 1.17; 95%CI 0.82-1.67) though confidence intervals overlapped. The sensitivity analysis comparing to the 0-13 day post dose 1 period gave a similar pattern of results though the odds ratios were smaller and not statistically significant (supplementary table 3). This was also the case with the matched analysis (supplementary table 4).

**Table 3:** Odds ratios for detection of B.1.617.2 relative to B.1.1.7 in vaccinated compared to unvaccinated individuals.

| Vaccination status | Number of cases |  | Ratio B.1.617.2 to B.1.1.7 | aOR |
| --- | --- | --- | --- | --- |
|  | B.1.1.7 | B.1.617.2 |  |  |
| Unvaccinated | 8268 | 691 | 0.084 | base |
| Any vaccine |  |  |  |  |
| Dose 1 | 2237 | 272 | 0.122 | 1.38 (1.10-1.72) |
| Dose 2 | 81 | 25 | 0.309 | 1.60 (0.87-2.97) |
| Dose 1 or 2 | 2511 | 322 | 0.128 | 1.40 (1.13-1.75) |
| Vaccine type (dose 1 or 2) |  |  |  |  |
| BNT162b2 | 720 | 68 | 0.094 | 1.17 (0.82-1.67) |
| ChAdOx1 | 1791 | 254 | 0.142 | 1.48 (1.18-1.87) |

aOR = Adjusted odds ratio – adjusted for: week of test and region (as linear interaction), history of travel, ethnicity, age, sex, clinically extremely vulnerable, care home resident

## Discussion

### Main findings

We found an absolute reduction of one dose vaccine effectiveness against symptomatic disease with the B.1.617.2 variant of approximately 20% when compared to the B.1.1.7 variant. However, reductions in vaccine effectiveness after two doses were very small. This was the case for both the BNT162b2 and ChAdOx1 vaccines. Using a TNCC analysis, estimated vaccine effectiveness against symptomatic disease with B.1.617.2 for a single dose of either vaccine is approximately 33%, for two doses of BNT162b2 is approximately 88% and for two doses of ChAdOx1 is approximately 60%.

### Interpretation

These findings suggest a modest reduction in vaccine effectiveness. Nevertheless, a clear effect of both vaccines was noted with high levels of effectiveness after two doses. Vaccine effects after two doses of ChAdOx1 vaccine were smaller than for BNT162b2 against either variant. This is consistent with reported clinical trial findings. However, rollout of second doses of ChAdOx1 was later than BNT162b2 and the difference may be explained by the limited follow-up after two doses of ChAdOx1 if it takes more than two weeks to reach maximum effectiveness with this vaccine. Consistent with this, 74% of those who had received 2 doses of ChAdOx1 had done so between 2 and 4 weeks prior to symptom onset compared to 46% with BNT162b2 (supplementary figure 1).

Numbers of cases and follow-up periods are currently insufficient to estimate effectiveness against severe disease, including hospitalisation and mortality, however, previous vaccine effectiveness estimates with other variants have shown higher levels of effectiveness against more severe outcomes.(10, 14, 28) Therefore higher levels of effectiveness against severe disease may be anticipated with the B.1.617.2 vaccine.

### Comparison with other studies

This is the first study that we are aware of to report on vaccine effectiveness against the B.1.617.2 variant. We were also unable to find any neutralisation data for this variant. One study from India has reported neutralisation data with the broader B.1.617 variant category suggests that both convalescent sera of COVID-19 cases and sera from recipients of the BBV152 (Covaxin) vaccine were able to neutralise B.1.617.(29) Assuming that a significant proportion of these were B.1.617.2 and that effectiveness the impact on different vaccines is similar, this would support our findings. Compared to recent findings from Qatar comparing the effectiveness of BNT162b2 against the B.1.1.7 and B1.351 variants, our findings would suggest that effectiveness against B.1.617.2 after a full course lies somewhere between these two.(17)

### Strengths and limitations

The large scale of testing and whole genome sequencing in the UK as well as the recording of vaccination status in a national vaccination register has allowed us to analyse vaccine effectiveness within a few weeks of the variant first emerging in the UK. We use two distinct analytical approaches which give broadly similar results and findings with our control analysis (using B.1.1.7) are consistent with those previously reported.(7, 8, 10, 17) Findings were also similar when comparing to the first 2 weeks post the first dose of vaccine (supplement), which helps to exclude unmeasured confounders associated with both the likelihood of being vaccinated and the likelihood of being exposed to a variant. Using a TNCC design also helped us to control for differences in health seeking behaviour between vaccinated and unvaccinated individuals.

There are also limitations to this study. These are observational findings and should be interpreted with caution. There may be factors that could increase the risk of COVID-19 in vaccinated individuals, for example if they adopt more risky behaviours following vaccination, however, this would be likely to affect analysis of both variants. Low sensitivity or specificity of PCR testing could also result in cases and controls being misclassified which would attenuate vaccine effectiveness estimates. This could affect one variant more than another, though this might be expected to affect B.1.1.7 more than B.1.617.2 as with an emerging variant more cases may be detected earlier in infection which may result in higher viral loads and increased sensitivity and specificity. While we control for ethnicity, region and level of deprivation, differences in vaccine coverage in population groups that may have more or less exposure to B.1.617.2 may have affected our first analysis, but should not have affected the TNCC design. There may also be differences in the populations that received each vaccine, for example among younger age groups, more healthcare workers are likely to have received BNT162b2 whereas more individuals in clinical risk groups are likely to have received ChAdOx1, while we control for these factors in the analysis, we cannot exclude residual confounding.(11) Timing of rollout of different vaccines varied, the reduced follow-up post two doses of ChAdOx1 could have attenuated these results, as discussed above. Furthermore, numbers who had received the Moderna mRNA-1273 vaccine were too small to be able to estimate vaccine effectiveness for this vaccine. As such, it is important that these findings are triangulated with emerging in vitro data on immune response in vaccinated individuals.

## Conclusions

Overall, we found high levels of vaccine effectiveness against symptomatic disease after two doses. These estimates were only modestly lower than vaccine effectiveness against the B.1.1.7 variant. It is likely that vaccine effectiveness against more severe disease outcomes will be greater. Our finding of reduced effectiveness after dose 1, would support maximising vaccine uptake with two doses among vulnerable groups in the context of circulation of B.1.617.2.

## Data Availability

All relevant data are available in the manuscript or supplement

## Statements

## Acknowledgements

We thank the Public Health England covid-19 Data Science Team,NHS England, NHS Digital, and NHS Test and Trace for their roles in developing and managing the covid-19 testing and vaccination systems and datasets as well as reporting NHS vaccinators, NHS laboratories, PHE laboratories, and lighthouse laboratories; and we thank the Wellcome Sanger Institute and other laboratories involved in whole genome sequencing of COVID-19 samples; and we thank the Joint Committee on Vaccination and Immunisation, the UK Variant Technical Group and the UK covid-19 Vaccine Effectiveness Working Group for advice and feedback in developing this study.

## Ethical approval

Surveillance of covid-19 testing and vaccination is undertaken under Regulation 3 of The Health Service (Control of Patient Information) Regulations 2002 to collect confidential patient information (www.legislation.gov.uk/uksi/2002/1438/regulation/3/made) under Sections 3(i) (a) to (c), 3(i)(d) (i) and (ii) and 3(3). The study protocol was subject to an internal review by the Public Health England Research Ethics and Governance Group and was found to be fully compliant with all regulatory requirements. As no regulatory issues were identified, and ethical review is not a requirement for this type of work, it was decided that a full ethical review would not be necessary.

