## supplementary for "Effectiveness of COVID-19 vaccines against the B.1.617.2 variant"

This preprint reports new research that has not been certified by peer review

### Supplementary appendix

*Supplementary table 1: Cross-tabulation of S-gene target status and variant as identified by whole genome sequencing*

| Variant | S-gene | | | Total |
|---|---|---|---|---|
| | positive | negative | unknown | |
| No sequencing result | 356 | 1,574 | 439 | 2,369 |
| B.1.1.7 | 2 | 5,099 | 278 | 5,379 |
| B.1.1351 | 19 | 0 | 4 | 23 |
| B.1.617.2 | 666 | 0 | 56 | 722 |
| P.2 | 7 | 0 | 0 | 7 |
| B.1.617.1 | 28 | 0 | 9 | 37 |
| B.1.617.3 | 3 | 0 | 0 | 3 |
| B.1.525 | 0 | 12 | 0 | 12 |
| B.1.1.318 | 9 | 0 | 1 | 10 |
| Low quality (likely B.1.617.2) | 1 | 0 | 1 | 2 |
| Not VOC/VUI | 26 | 3 | 3 | 32 |
| Total | 1,117 | 6,688 | 791 | 118,157 |

*Supplementary table 2: Cross-tabulation of S-gene target status and variant as identified by whole genome sequencing after dropping non B.1.1.7 or B.1.617.2 variants and reassignment based on sequencing results*

| Variant | S-gene | | | Total |
|---|---|---|---|---|
| | positive | negative | unknown | |
| No sequencing result | 356 | 1,574 | 439 | 2,369 |
| B.1.1.7 | 0 | 5,379 | 0 | 5,379 |
| B.1.617.2 | 722 | 0 | 0 | 722 |
| Total | 1,117 | 6,688 | 791 | 118,157 |

Preprint

This preprint reports new research that has not been certified by peer review

*Supplementary table 3: Odds ratios for detection of B.1.617.2 relative to B.1.1.7 in vaccinated individuals compared to the <14 days post dose 1*

| Vaccination status | Number of cases | | Ratio B.1.617.2 to B.1.1.7 | aOR |
|---|---|---|---|---|
| | B.1.1.7 | B.1.617.2 | | |
| Days 0-13 post dose 1 | 551 | 32 | 0.058 | base |
| Any vaccine | | | | |
| Dose 1 | 2237 | 272 | 0.122 | 1.30 (0.82-2.08) |
| Dose 2 | 81 | 25 | 0.309 | 1.52 (0.72-3.18) |
| Dose 1 or 2 | 2511 | 322 | 0.128 | 1.33 (0.83-2.11) |
| Vaccine type (dose 1 or 2) | | | | |
| BNT162b2 | 720 | 68 | 0.094 | 1.10 (0.64-1.90) |
| ChAdOx1 | 1791 | 254 | 0.142 | 1.39 (0.87-2.23) |

*Supplementary table 4: Matched case control analysis*

| Vaccination status | Number of cases | | Ratio B.1.617.2 to B.1.1.7 | aOR compared to unvaccinated | aOR compared to <14 days post dose |
|---|---|---|---|---|---|
| | B.1.1.7 | B.1.617.2 | | | |
| Unvaccinated | 2799 | 534 | 0.191 | base | |
| <14 days post dose 1 | 133 | 21 | 0.158 | | base |
| Any vaccine | | | | | |
| Dose 1 | 621 | 173 | 0.279 | 1.19 (0.93-1.54) | 1.29 (0.74-2.22) |
| Dose 2 | 33 | 13 | 0.394 | 1.44 (0.70-2.95) | 1.55 (0.65-3.70) |
| Dose 1 or 2 | 710 | 198 | 0.279 | 1.18 (0.93-1.51) | 1.27 (0.74-2.19) |

Matched on Ethnicity, Region, age(10 yrs), week of sample

Preprint

This preprint reports new research that has not been certified by peer review

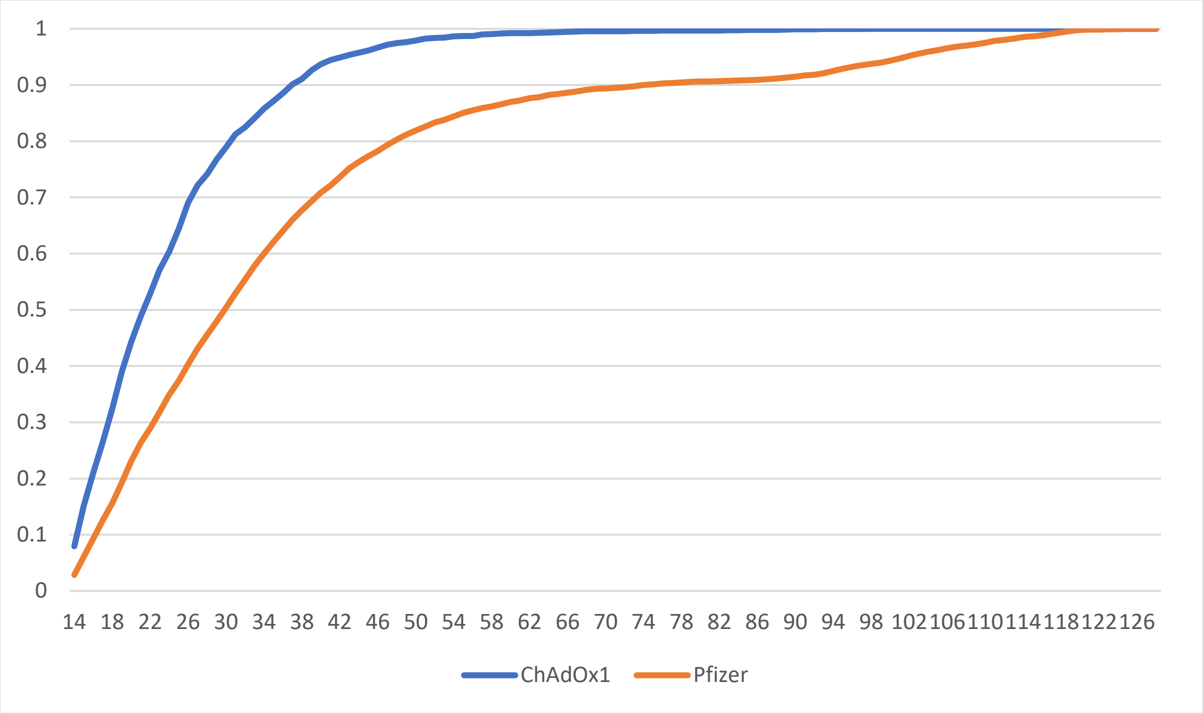

*Figure 1: Cumulative proportion of those with 2 doses more than 14 days after dose 2 vaccinated by each time point after the second dose*

Preprint
